# Improving hand hygiene in community settings: a scoping review of current international guidelines

**DOI:** 10.1101/2022.09.29.22280518

**Authors:** Clara MacLeod, Laura Braun, Bethany A. Caruso, Claire Chase, Kondwani Chidziwisano, Jenala Chipungu, Robert Dreibelbis, Regina Ejemot-Nwadiaro, Bruce Gordon, Joanna Esteves Mills, Oliver Cumming

## Abstract

**Background:** Hand hygiene is an important measure to prevent disease transmission in community settings, such as households, public spaces, workplaces, and schools. There exist various international guidelines with recommendations on how to improve hand hygiene in these settings, but no review to date has been conducted to summarise these recommendations and assess to what extent they are consistent and evidence-based.

**Methods:** To identify international guidelines with recommendations on hand hygiene in community settings, categorised as either domestic, public, and institutional, we performed electronic and grey literature searches and contacted expert organisations and individuals. Recommendations extracted from included guidelines were mapped to four areas related to hand hygiene: i) effective hand hygiene; ii) minimum requirements; iii) behaviour change; and iv) government measures. We assessed if recommendations were supported by peer-reviewed literature and checked their consistency and concordance across settings.

**Results:** We identified 51 guidelines published between 1999 and 2021 by multilateral agencies and international non-governmental organisations containing 923 recommendations. Handwashing with soap is consistently recommended as the preferred method for hand hygiene across all community settings. Most guidelines specifically recommend handwashing with plain soap and running water for at least 20 seconds; single-use paper towels for hand drying; and alcohol-based hand rub (ABHR) as a complement or alternative to handwashing. There are inconsistent and discordant recommendations for water quality for handwashing, affordable and effective alternatives to soap and ABHR, and the design of handwashing stations. Further, there are gaps in recommendations on soap and water quantity, behaviour change approaches, and government measures required for effective hand hygiene. Overall, less than 10% of recommendations are supported by evidence.

**Conclusion:** While current international guidelines consistently recommend handwashing with soap in domestic, public, and institutional settings, the lack of consistent, evidence-based recommendations may constrain global efforts to ensure effective hand hygiene across community settings.

**KEY MESSAGES:** *What is already known on this topic:*

- Hand hygiene has been found to be a cost-effective intervention that can reduce the risk of certain infectious diseases.
- Yet, the practice of hand hygiene, and access to the facilities which enable this, is often limited in community settings, such as households, public spaces, workplaces, and schools.
- There are various international guidelines with recommendations on hand hygiene in community settings, but it is unclear whether guidelines provide consistent and evidence-based recommendations.

*What this study adds:*

- There are 51 guidelines with over 900 recommendations for hand hygiene in community settings published by multilateral agencies and international non-governmental organisations.
- Guidelines consistently recommend handwashing with soap, but there are several areas of inconsistency and discordance, as well as gaps in recommendations, related to minimum requirements, behaviour change, and government measures for effective hand hygiene in community settings.
- Very few recommendations are supported by peer-reviewed literature.

*How this study might affect research, practice, or policy:*

- This scoping review highlights a gap in global normative guidance on hand hygiene in community settings.
- More research is needed to address the current areas of inconsistency and discordance, and gaps in recommendations.

## INTRODUCTION

Hand hygiene, including handwashing with soap and other methods such as alcohol-based hand rubs (ABHRs), is an important public health measure that can prevent the transmission of a range of diseases.^1^ Handwashing with soap has been found to be a cost-effective intervention^2^ that can reduce the risk of both diarrhoeal disease and acute respiratory infections (ARIs) by over 20%.^3–9^ Handwashing with soap has also been linked to the reduction of certain neglected tropical diseases (NTDs), including trachoma and some soil-transmitted helminth infections.^10,11^ Recently, handwashing with soap and the use of ABHRs were advised as one of the key control measures during the COVID-19 pandemic^12,13^ and were found to be effective.^14^

This scoping review focuses on hand hygiene in non-healthcare settings, which we collectively refer to as “community settings”. Using the definition set out in the Ottawa Charter, we consider settings as where “health is created and lived by people within the settings of their everyday life; where they learn, work, play and love”,^15^ and include domestic, public, and institutional settings. The practice of hand hygiene – and access to the facilities which enable this – is often limited in these community settings, particularly in low- and middle-income countries.^16^ In the domestic setting, 30% of the global population does not have access to a basic handwashing facility with soap and water at home,^17,18^ with three quarters of those living in low-income countries.^17^ In institutional settings, an estimated 43% of schools worldwide do not have access to basic hand hygiene facilities^18^ but there is limited data for other institutional settings, such as the workplace and prisons and places of detention, and public settings, such as markets, transportation hubs, and places of worship.^18^

Despite the international recognition of hand hygiene as a critical public health measure, a recent global assessment of government policies, planning, and financing for hygiene found that while the majority of surveyed countries reported having national policies for hand hygiene, less than 10% had sufficient financing to implement them.^19^ Various international guidelines with recommendations on hand hygiene for non-healthcare settings exist^13,20–22^ but it is unclear whether current international guidelines are comprehensive, consistent, and based on the most rigorous evidence available. This review aims to summarise current international recommendations for hand hygiene in community settings, identify areas of consistency and concordance, and assess whether recommendations are evidence-based.

## METHODS

This review follows the six stages of the Arksey and O’Malley methodological framework for scoping reviews.^23–25^ Expert consultation (stage 6) was integrated throughout the scoping review process^24^ to obtain feedback on the scope and conceptual framework and to identify any additional guidelines beyond those identified during the electronic search. Our review is described according to the Preferred Reporting Items for Systematic reviews and Meta-Analyses extension for Scoping Reviews (PRISMA-ScR)^26^ and a PRISMA-ScR checklist is included in the supplementary materials. (Table A1). The protocol was pre-registered with OSF Registries.^27^

### Identifying the research question (stage 1)

To identify and refine the research question, a conceptual framework was developed for this review and built around three key concepts: i) hand hygiene, ii) non-healthcare settings, and iii) current international guidelines (Figure 1). In this review, we define hand hygiene as any action of hand cleansing for the purpose of removing or deactivating pathogens from hands.^28^ Effective hand hygiene is defined as any practice which removes or deactivates pathogens from hands and thereby limits disease transmission.^28^

**Figure 1.**
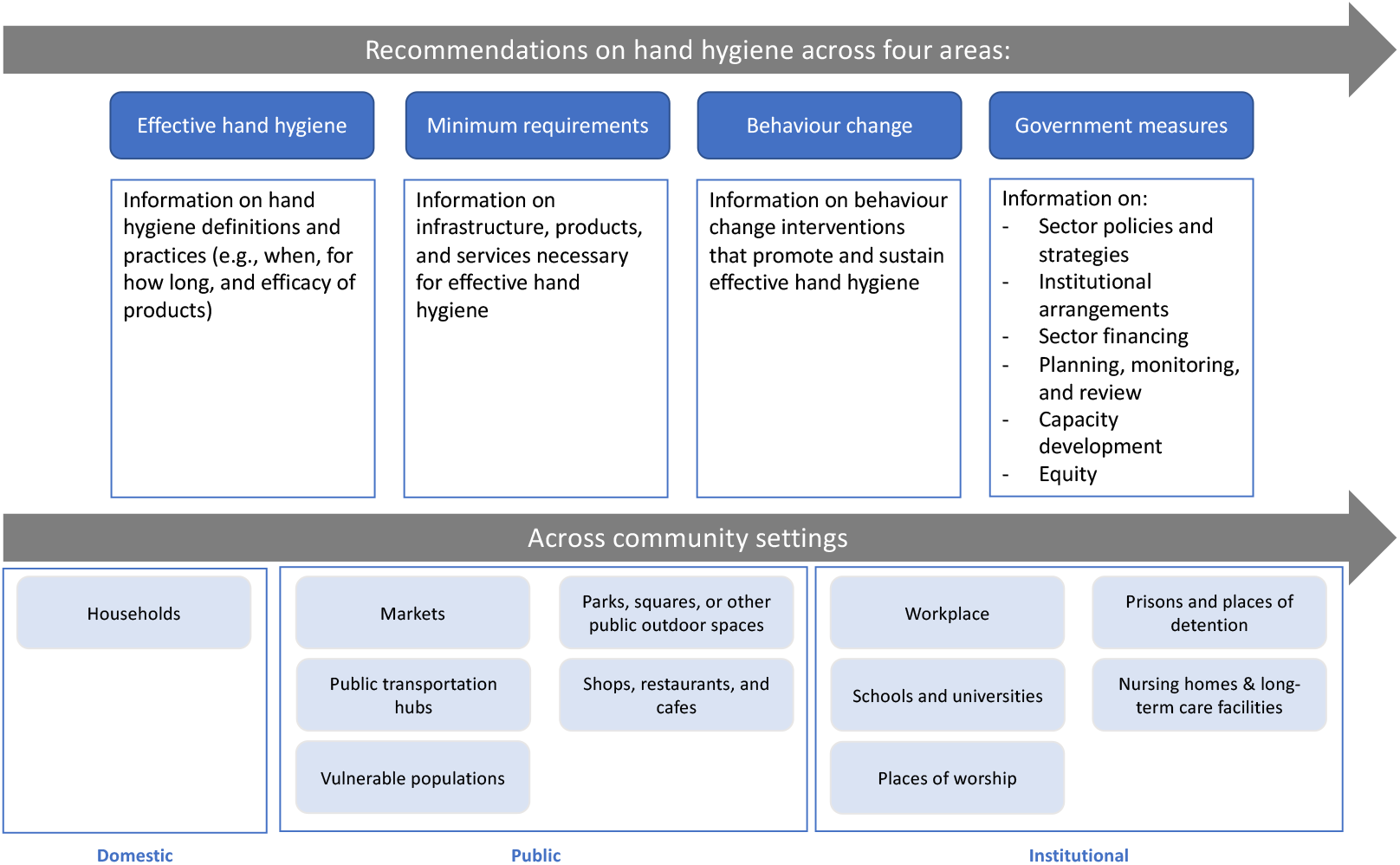
Conceptual framework for hand hygiene in community settings.

The first key concept in the conceptual framework – hand hygiene – covers four areas: effective hand hygiene, minimum requirements, behaviour change, and government measures (Figure 1). Effective hand hygiene refers to definitions and practices. Minimum requirements refer to the materials, services, and infrastructure required for effective hand hygiene. Behaviour change denotes the appropriate behavioural performances that promote and sustain effective hand hygiene. Government measures concern actions taken by governments to ensure effective hand hygiene, which are categorised according to an established framework^29^ as follows: policy and strategy; institutional arrangements; sector financing; planning, monitoring, and review; capacity development; and equity.

The second key concept – community settings – is defined as settings where healthcare is not routinely delivered,^28^ broadly spanning all places where people “learn, play, work and love”^15^ and specifically including domestic, public, and institutional settings. As these settings may not exclusively refer to physical settings,^30^ the review also includes recommendations for vulnerable populations (e.g., people experiencing homelessness) who may reside permanently or semi-permanently in public spaces. Nursing homes and long-term care facilities are also included in the review under institutional settings, as the boundary between healthcare and non-healthcare settings is often less clear.

The third concept – guideline – is defined as a published document where the primary purpose is to provide specific guidance, in the form of recommendations, towards a course of action. A recommendation is defined as a statement designed to assist a targeted actor to take informed decisions on whether, when, and how to undertake a specific action.^31^ Our review is limited to international guidelines to identify generalisable recommendations of global relevance.

### Identifying relevant studies (stage 2)

To identify relevant guidelines, we searched the WHO Institutional Repository for Information Sharing (IRIS) database and grey literature (Google search engine and websites of international organisations known to work on hand hygiene) (Table A2) using pre-specified search terms related to hand hygiene, non-healthcare settings, and guidelines (Table A3). Expert consultations were also conducted with ‘Hand Hygiene for All’ Initiative core partners (Table A4) to identify potentially relevant guidelines and reference lists of guidelines were hand-searched. The search was limited to English and French languages and publication date was restricted to 1990 onwards to identify current guidelines.

### Study selection (stage 3)

Documents meeting the following criteria were included: i) international guideline, ii) offers one or more recommendations on hand hygiene, iii) targets one or more community setting, as defined in the conceptual framework, iv) published by an international NGO, multilateral agency, or public health agency, v) published in English or French, and vi) published between 1 January 1990 and 1 November 2021. The review excludes guidelines for humanitarian settings, as internationally agreed guidance on hand hygiene in humanitarian settings and complex emergencies is available through the Sphere standards for water, sanitation, and hygiene promotion.^32^ Only the most recent versions of guidelines were included, with previous versions of the same guidelines excluded. Country-specific guidelines were also excluded.

All documents retrieved from electronic searches and expert consultations were transferred to Mendeley^33^ for de-duplication. Inclusion was completed in two stages: (1) title and abstracts were screened for eligibility by one reviewer (CM); and (2) then full text for all potentially eligible documents were retrieved and independently assessed for inclusion by two reviewers (CM and LB). Disagreement between reviewers on inclusion was resolved through arbitration by a third reviewer (OC).

### Charting the data (stage 4)

Guideline characteristics and recommendations from included guidelines were double-extracted by two reviewers (CM and LB) using a standardised data extraction template in MS Excel^34^ and then cross-checked for accuracy. As with inclusion, a third reviewer (OC) provided arbitration if agreement on extraction could not be reached. The data extraction form included information on guideline characteristics, such as author, year of publication, target setting, and COVID-19 response, as well as 57 specific parameters related to the four areas of hand hygiene described in the conceptual framework (Figure 1). Recommendations for each parameter were extracted from included guidelines where possible.

### Collating, summarising, and reporting the results (stage 5)

Guideline recommendations were first summarised for each parameter across community settings and then disaggregated by domestic, public, or institutional setting where relevant. Definitions and recommendations for hand hygiene were assessed for consistency, concordance, and whether supported by evidence. Recommendations were classified as consistent, fairly consistent, or inconsistent if they featured in 10 or more, four to nine, or less than four guidelines, respectively. Concordance is here defined as parameters with no consistent or fairly consistent recommendations at odds with each other. We also assessed whether extracted recommendations were supported by one of four types of evidence, including peer-reviewed literature, grey literature, other guideline, or other sources (e.g., programme documentation). A recommendation was considered evidence-based if the guideline provided a specific citation for the recommendation. Each evidence-based recommendation was then coded according to the type of evidence cited. Finally, evidence gaps were defined as parameters with very few recommendations (i.e., less than 10 recommendations, equivalent to less than 20% of guidelines providing a recommendation).

### Patient and public involvement

There was no public or patient involvement in the course of this project.

## RESULTS

### Search results

Electronic searches were conducted on 15 November 2021 identifying 2,492 records (2,432 from the WHO IRIS database, 32 from Google, 28 from international agency websites) (Table A2) and a further 11 records identified through expert consultation. Following de-duplication, a total of 2,264 records were screened by title and abstract, and 125 documents were sought for retrieval for full-text screening, with one document not accessible. Finally, 51 guidelines are included in the review (Figure 2). The 73 documents excluded during the full-text review are listed in the Appendix with reasons for exclusion (Table A5).

**Figure 2.**
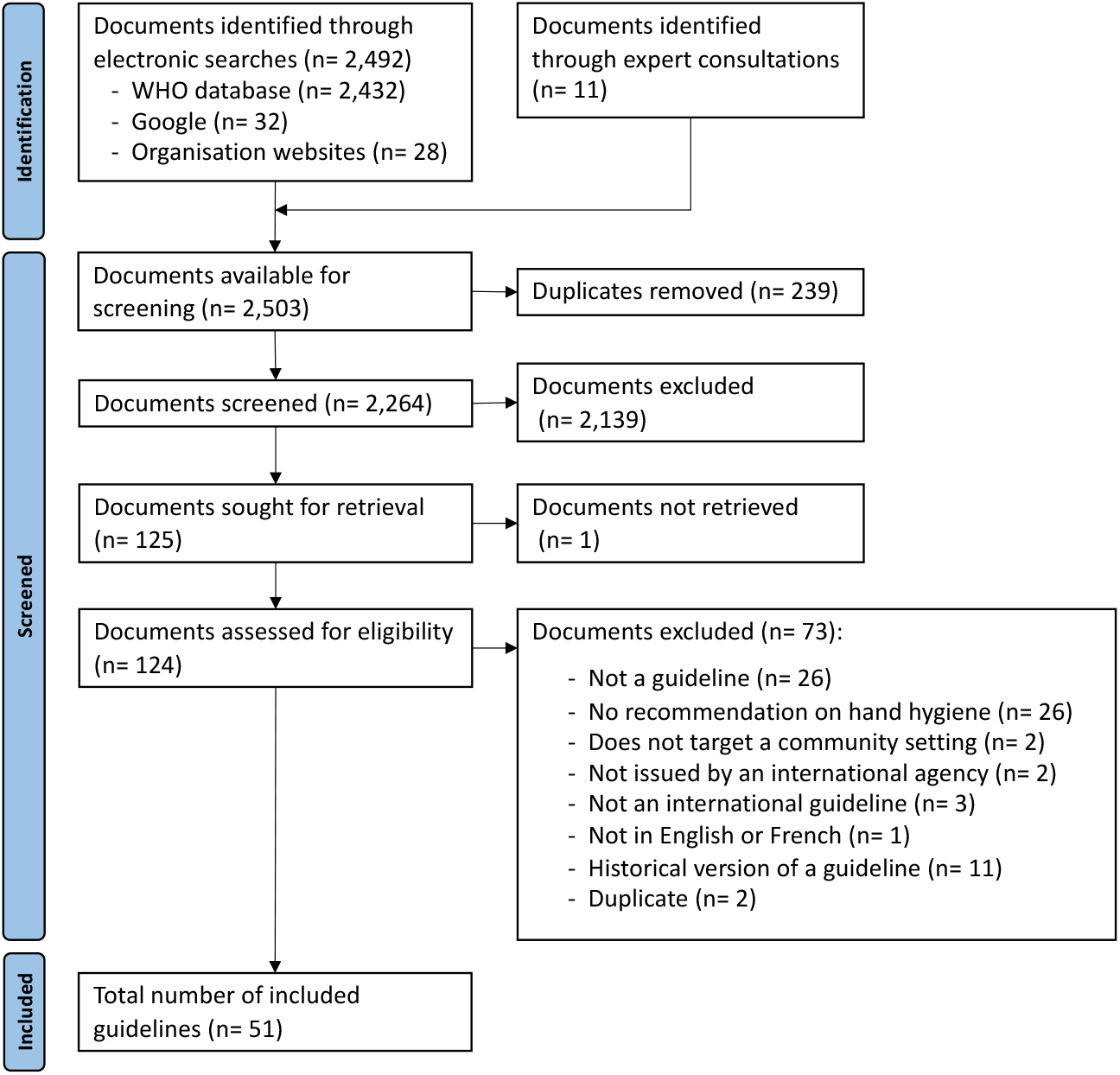
PRISMA flow diagram.

### Description of included guidelines

The 51 included guidelines were published in English between 1999 and 2021, with 38 published in 2020 or later and 31 providing COVID-19-specific guidance. Among the 51 included guidelines, 67% are published by multilateral agencies (WHO, UNICEF, UNHCR, and ILO), 23% by international NGOs, and 10% by the United States Centers for Disease Control and Prevention (US CDC) (Table A6). Most guidelines target public and institutional settings, while none exclusively target the domestic setting.

More specifically, 43% (n= 22) target the public setting, 43% (n= 22) the institutional setting, 4% (n= 2) the domestic and public setting, and 10% (n= 5) more than one setting (e.g., domestic, public, and/or institutional). Of the 22 guidelines for the public setting, 20 concern public spaces, and two concern vulnerable populations within public spaces (e.g., people experiencing unsheltered homelessness and people living in dense, informal settlements). For institutional settings, eight guidelines concern schools, six the workplace, four prisons and places of detention, three places of worship, and one long-term care facilities. Almost no guidelines, however, define a community setting. Overall, we extracted 923 recommendations from the 51 included guidelines (Table A7).

### Recommendations for effective hand hygiene

#### Hand hygiene definitions

Only 10% of guidelines provide a clear definition for hand hygiene. Meanwhile, 75% of guidelines provide at least one recommendation on when to practice hand hygiene, referred to in our review as a ‘key moment’. Different terms are used for key moments including ‘key times’ (14%), ‘critical times’ (8%), and ‘key moments’ (4%). Otherwise, guidelines either do not use a specific term for defining a key moment (49%) or do not recommend at least one key moment (25%).

#### Hand hygiene practices

There is agreement among guidelines on what constitutes effective hand hygiene across community settings, although there are gaps in recommendations on how to practice effective hand hygiene. Almost all guidelines (90%) recommend washing hands with soap and for a duration of at least 20 seconds (27%) (Table 1). In addition, 63% of guidelines recommend the use of ABHR as an alternative or complement to handwashing with soap, though very few guidelines recommend a duration for hand rubbing. Few guidelines provide specific recommendations on handwashing or hand rubbing technique (14%), with over half of these guidelines (60%, n= 4) referring to the WHO instructions for hand hygiene in healthcare settings.^28^ Meanwhile, 24% of guidelines recommend including simple instructions on handwashing technique in hygiene promotion programmes.

**Table 1.** Consistent and fairly consistent recommendations for hand hygiene in community settings.

| Parameter | Recommendation | All guidelines (n) | Domestic setting (n) | Public setting (n) | Institutional setting (n) | Multiple settings (n) |
| --- | --- | --- | --- | --- | --- | --- |
| Number of guidelines |  | 51 | 2 | 24 | 22 | 5 |
| **Effective hand hygiene** |  |  |  |  |  |  |
| Key moments | Before and after eating | 45% (23) | 0% (0) | 38% (9) | 27% (14) | 0% (0) |
|  | After using the toilet | 41% (21) | 0% (0) | 38% (9) | 55% (12) | 0% (0) |
|  | Before and after preparing food | 27% (14) | 0% (0) | 38% (9) | 41% (9) | 20% (1) |
|  | After blowing nose, coughing, or sneezing | 24% (12) | 0% (0) | 21% (5) | 27% (6) | 20% (1) |
|  | After touching public surfaces or objects | 24% (12) | 0% (0) | 12% (6) | 18% (4) | 40% (2) |
|  | When entering/exiting buildings or home | 14% (7) | 0% (0) | 17% (4) | 5% (1) | 40% (2) |
|  | After changing a child's diaper | 12% (6) | 0% (0) | 17% (4) | 0% (0) | 40% (2) |
|  | When hands visibly dirty | 10% (5) | 0% (0) | 13% (3) | 9% (2) | 0% (0) |
|  | Before and after work | 8% (4) | 0% (0) | 0% (0) | 18% (4) | 0% (0) |
|  | After contact with animals | 8% (4) | 0% (0) | 13% (3) | 5% (1) | 0% (0) |
|  | Taking care of sick person | 8% (4) | 0% (0) | 13% (3) | 5% (1) | 0% (0) |
| Handwashing duration | At least 20 seconds | 27% (14) | 0% (0) | 21% (5) | 36% (8) | 20% (1) |
| **Minimum requirements for effective hand hygiene** |  |  |  |  |  |  |
| **Materials for effective hand hygiene** |  |  |  |  |  |  |
| Soap | Soap | 65% (33) | 100% (2) | 50% (12) | 82% (18) | 60% (3) |
|  | Bar soap | 24% (12) | 0% (0) | 14% (7) | 14% (3) | 40% (2) |
|  | Liquid soap | 22% (11) | 0% (0) | 14% (7) | 14% (3) | 20% (1) |
|  | Soap water (e.g., powder, liquid, or bar soap shavings diluted in water) | 16% (8) | 0% (0) | 17% (4) | 5% (1) | 60% (3) |
| ABHR | ABHR with at least 60% alcohol | 35% (18) | 0% (0) | 16% (8) | 45% (10) | 0% (0) |
|  | ABHR, no alcohol percentage | 22% (11) | 50% (1) | 17% (4) | 23% (5) | 20% (1) |
| Hand drying | Clean, single use paper towels | 39% (20) | 0% (0) | 14% (7) | 59% (13) | 0% (0) |
|  | Air-drying system | 22% (11) | 0% (0) | 17% (4) | 14% (7) | 0% (0) |
|  | Bin with disposable liners and lid | 8% (4) | 0% (0) | 4% (1) | 14% (3) | 0% (0) |
| **Alternative materials** |  |  |  |  |  |  |
| Other materials | Ash | 22% (11) | 0% (0) | 12% (6) | 14% (3) | 40% (2) |
| Conditional recommendations | ABHR where soap and water not available | 18% (9) | 0% (0) | 4% (1) | 36% (8) | 0% (0) |
|  | Ash and water if ABHR or soap not available | 14% (7) | 0% (0) | 13% (3) | 14% (3) | 20% (1) |
|  | ABHR if hands not visibly soiled | 10% (5) | 0% (0) | 8% (2) | 14% (3) | 0% (0) |
| **Water requirements** |  |  |  |  |  |  |
| Water services | Running water | 43% (22) | 0% (0) | 16% (8) | 55% (12) | 40% (2) |
|  | Piped water system connected to tap | 16% (8) | 0% (0) | 13% (3) | 18% (4) | 20% (1) |
|  | Water trucking, storage, or manual refilling | 14% (7) | 0% (0) | 13% (3) | 14% (3) | 20% (1) |
| Alternative water sources for handwashing | Rainwater | 8% (4) | 0% (0) | 17% (4) | 0% (0) | 0% (0) |
| Wastewater management | Drainage system | 16% (8) | 0% (0) | 8% (2) | 23% (5) | 20% (1) |
|  | Covered soakaway pit | 12% (6) | 0% (0) | 21% (5) | 0% (0) | 20% (1) |
| **Hand hygiene stations** |  |  |  |  |  |  |
| Handwashing station | Washbasin | 16% (8) | 0% (0) | 13% (3) | 18% (4) | 20% (1) |
|  | Bucket or container connected to tap | 14% (7) | 50% (1) | 17% (4) | 5% (1) | 40% (2) |
|  | Tippy tap | 12% (6) | 0% (0) | 4% (1) | 14% (3) | 40% (2) |
|  | Foot-pedal or hand-operated system | 8% (4) | 0% (0) | 8% (2) | 5% (1) | 20% (1) |
| Location of handwashing stations | Key entry/exit points | (14) | 0% (0) | 50% (12) | 5% (1) | 20% (1) |
|  | Close proximity to toilets | (14) | 0% (0) | 21% (5) | 14% (7) | 40% (2) |
|  | food preparation and eating areas | (9) | 0% (0) | 8% (2) | 27% (6) | 20% (1) |
| Accessibility | Accessible for all users and vulnerable groups | 12% (6) | 0% (0) | 13% (3) | 5% (1) | 40% (2) |
|  | Height of soap and water taps appropriate for children, elderly, and disabled | 10% (5) | 0% (0) | 13% (3) | 5% (1) | 20% (1) |
|  | Age-appropriate handwashing stations | 8% (4) | 0% (0) | 0% (0) | 18% (4) | 0% (0) |
| Design considerations | Theft resistance | 16% (8) | 0% (0) | 17% (4) | 14% (3) | 20% (1) |
|  | Water-saving designs/technologies | 12% (6) | 0% (0) | 13% (3) | 9% (2) | 20% (1) |
| COVID-19 adaptations | Taps that limit cross-contamination | 16% (8) | 0% (0) | 14% (7) | 0% (0) | 20% (1) |
|  | 1-2m spacing between handwashing stations | 12% (6) | 0% (0) | 17% (4) | 5% (1) | 20% (1) |
|  | Disinfect taps regularly | 10% (5) | 0% (0) | 17% (4) | 0% (0) | 20% (1) |
|  | Towels for opening/closing taps | 8% (4) | 0% (0) | 13% (3) | 5% (1) | 0% (0) |
| Availability of materials | Locally available materials for handwashing | 10% (5) | 0% (0) | 8% (2) | 9% (2) | 20% (1) |
| **Behaviour change** |  |  |  |  |  |  |
| Behaviour change techniques<sup>1</sup> | Prompts and cues | 12% (6) | 0% (0) | 13% (3) | 14% (3) | 0% (0) |
|  | Habit formation | 8% (4) | 0% (0) | 13% (3) | 5% (1) | 0% (0) |
| Content of behaviour change messaging | Doable instructions with proper steps | 24% (12) | 0% (0) | 12% (6) | 27% (6) | 0% (0) |
|  | Messages that target motivation | 10% (5) | 0% (0) | 8% (2) | 14% (3) | 0% (0) |
| Delivery channels | Visual reminders | 31% (16) | 0% (0) | 12% (6) | 36% (8) | 40% (2) |
|  | Mass communication | 24% (12) | 0% (0) | 14% (7) | 9% (2) | 60% (3) |
|  | Small group activities | 18% (9) | 0% (0) | 12% (6) | 9% (2) | 20% (1) |
|  | Interpersonal communication | 10% (5) | 0% (0) | 17% (4) | 0% (0) | 20% (1) |
| Formative research | Identify determinants of target behaviour within target population | 8% (4) | 0% (0) | 4% (1) | 0% (0) | 60% (3) |

<sup>1</sup> Using a typology of behaviour change techniques developed by Michie et al. (2013)<sup>36</sup>

Guidelines provide inconsistent recommendations on when to practice hand hygiene. 75% (n= 38) of guidelines specify a key moment for hand hygiene, though over 30 different individual key moments are recommended across guidelines. The most common individual key moments include before and after eating, after using the toilet, before and after preparing food, after blowing nose, coughing, or sneezing, and after touching public surfaces or objects (Table 1). The latter two feature most commonly among guidelines published during the COVID-19 pandemic. Fairly consistently recommended individual key moments include when entering or exiting the home or public space, after changing a child’s diaper, and when hands are visibly dirty. However, almost all guidelines recommend clusters of key moments (i.e., individual key moments recommended together). The most common ones are ‘after using the toilet’ with ‘before and after eating’ (39%, n= 20); ‘after using the toilet’ with ‘before and after preparing food’ (24%, n= 12); and ‘after using the toilet’, with ‘before and after eating’, and ‘after blowing nose, coughing, or sneezing’ (22%, n= 11).

### Recommendations for minimum requirements

#### Handwashing materials

Guidelines consistently recommend plain soap for handwashing and paper towels or electric dryers for hand drying. Most guidelines (65%) do not recommend a specific type of soap for handwashing, though some specifically recommend bar soap (24%), liquid soap (22%), or “soapy water” (e.g., home-made mixture of powder or liquid soap, or bar soap shavings diluted in water) (16%) (Table 1). No guidelines specifically recommend antibacterial soap. For hand drying after handwashing, most guidelines recommend clean, single use paper towels (39%) and electric air-drying systems (22%). 8% of guidelines recommend a bin with disposal liners and lid for the waste management of paper towels.

#### Soap and water requirements for handwashing

While guidelines consistently recommend running water for handwashing with soap, there are gaps for soap and water quantity and discordant recommendations on water quality for handwashing. In terms of water services, almost half (43%) of guidelines recommend running water, 16% a piped water system connected to a tap, and 14% water trucking, storage, or manual refilling. Few guidelines recommend a minimum quantity of soap (14%) or water (18%) required for handwashing (Table 2). For water quality, 16% of guidelines state that water must be of drinking water quality in line with WHO guidelines.^35^ In contrast, 8% state that it does not have to be of drinking water quality, though none specify a quantitative standard nor whether non-drinking quality water may conditionally be used if high quality water is not available. In addition, 10% of guidelines recommend that free residual chlorine must be greater than or equal to 0.5 mg/l after at least 30 minutes of contact time.

**Table 2.** Parameters with gaps in recommendations (i.e., fewer than 10 recommendations, equivalent to less than 20% of guidelines providing a recommendation).

| Parameters with gaps in recommendations<sup>1</sup> | Percentage (%) of guidelines that provide a recommendation (n) |
| --- | --- |
| **Effective hand hygiene** |  |
| No definition of hand hygiene | 4% (2) |
| No definition of safe hand hygiene | 6% (3) |
| Handwashing knowledge | 16% (8) |
| Hand rubbing duration | 6% (3) |
| **Minimum requirements for effective hand hygiene** |  |
| **Soap and water requirements for handwashing** |  |
| Quantity of soap | 14% (7) |
| Quantity of water | 18% (9) |
| **Hand hygiene stations** |  |
| Soap supply | 6% (3) |
| Functionality | 10% (5) |
| Cost and affordability | 14% (7) |
| Durability | 16% (8) |
| Operation and maintenance – responsibility | 16% (8) |
| Operation and maintenance – actions | 18% (9) |
| Spacing and number of users per handwashing station | 18% (9) |
| **Behaviour and behaviour change** |  |
| Frequency of behaviour change interventions | 4% (2) |
| Behaviour change approaches | 18% (9) |
| **Government measures** |  |
| Sector policy and strategy | 0% (0) |
| Sector financing | 0% (0) |
| Capacity development | 2% (1) |
| Equity | 2% (1) |
| Planning, monitoring, and review | 4% (2) |
| Institutional arrangements | 10% (5) |

<sup>1</sup> Parameters with less than 20% of guidelines that provide a recommendation

#### Alternative materials for hand hygiene

There are consistent recommendations for ABHR as an alternative material for hand hygiene. Of the guidelines that recommend ABHR, 35% (n= 18) recommend ABHR with at least 60% alcohol, 22% (n= 11) do not specify an alcohol percentage, and only 6% (n= 3) recommend an alcohol percentage of at least 70%. In addition, 18% of guidelines recommend ABHR where soap and water are not available and 10% (n= 5) only if hands are not visibly soiled.

There are discordant recommendations for the use of ash. 22% of guidelines recommend ash as an alternative material to soap for handwashing. In addition, 14% recommend ash if ABHR or soap are not available. However, 14% of guidelines advise against the use of ash or other products, such as soil, sand, mud, or water alone. In addition, while some guidelines (8%) recommend 0.05% chlorine solution, one guideline advises against it.

When disaggregated by setting, ABHR is most consistently recommended in public and institutional settings where it may meet larger and more frequent demand than handwashing stations. Similarly, recommendations for the conditional use of ABHR where soap and water are not available feature the most in the institutional setting, particularly in the workplace and schools. Ash is most commonly recommended in the public setting, which includes low-resource and water-scarce settings where soap and water may not be available.

#### Hand hygiene stations

Overall, there are inconsistent recommendations on hand hygiene facilities and their location, as well as gaps. Guidelines recommend washbasins (e.g., ceramic, cement, or plastic) (16%), a bucket or container connected to tap (14%), and tippy taps (12%) for handwashing stations across all settings (Table 1). Guidelines specify 16 different locations for handwashing stations, which include by the entrance and exit of public spaces and buildings (e.g., restaurants, shops, markets, places of worship, train and bus stations) (27%), in close proximity to toilets (27%), and next to food preparation and eating areas (18%). Guidelines also mention placing hand hygiene stations, such as ABHR dispensers, at key entry and exit points of public spaces and buildings (14%). There are inconsistent recommendations on the optimal spacing and number of users per handwashing station and none for ABHR dispensers. Gaps in recommendations include those for hand hygiene materials and product supply chains, cost and affordability, functionality, durability, and hand hygiene station operation and maintenance responsibilities and actions (Table 2).

Recommendations on the location of handwashing stations vary slightly by setting. Close proximity to toilets is consistently recommended for both public and institutional settings (10% and 14%, respectively). By the entrance and exit of public spaces is most consistently recommended for public settings (24%). Next to food preparation and eating areas (12%) is most consistently recommended for institutional settings, such as schools and the workplace.

#### Hand hygiene station access and adaptations

Guidelines recommend hand hygiene stations that are accessible for all, including people with disabilities and older adults, and adapted for pandemic response. For example, 12% of guidelines recommend that hand hygiene stations be accessible for all users and vulnerable groups. In addition, 10% recommend that the height of soap and water taps be appropriate for access by children, the elderly, and disabled (e.g., 500-700 mm basin height for children and 850 mm basin height for wheelchair access) (Table 1). 45% (n= 23) of guidelines recommend COVID-19 adaptations for hand hygiene stations, such as taps that limit cross contamination (16%), one to two meters spacing between stations (12%), regular tap disinfection (10%), and towels for opening and closing taps (8%). Other adaptations include theft resistance (e.g., attaching soap or other movable pieces to the station) (16%) and water-saving designs (e.g., low-flow handwashing stations) (12%).

### Recommendations for behaviour change

Overall, there are few recommendations related to hand hygiene behaviour change, though there are consistent recommendations on behaviour change messaging and delivery channels. For behaviour change messaging, some guidelines (10%) recommend messages that target “motives”^37^ (Table 1). In terms of delivery channels, 31% of guidelines recommend using visual reminders (e.g., signs, posters, or leaflets) and 24% mass communication (e.g., radio, social media, or SMS) to deliver behaviour change messages. Prompts, cues and habit formation are fairly consistently recommended as behaviour change techniques.^36^ Formative research for behaviour change programmes is recommended by 10% (n= 5) of guidelines, though only 8% (n= 4) specifically mention undertaking formative research to identify behavioural determinants among the target population. Guidelines provide inconsistent recommendations on behaviour change approaches (18%), determinants of hand hygiene to target for interventions (22%), and behaviour change models or frameworks (33%). Lastly, there are gaps in recommendations on frequency (i.e., how often an intervention should be delivered) of behaviour change interventions and approaches (4%) (Table 2).

### Recommendations for government measures

Overall, few guidelines (22%, n= 11) provide a recommendation on government measures. Nonetheless, recommendations for sector policy and strategy include promoting local soap production and fostering public-private partnerships for handwashing (4%). For institutional arrangements, 4% of guidelines suggest identifying ways of cross-sectoral collaboration for hand hygiene, while other recommendations centre on engaging communities, the private sector, and civil society for the delivery of water, sanitation, and hygiene (WASH) services. On planning, monitoring, and review, 4% of guidelines recommend supporting or reinforcing existing monitoring systems or creating a government-led national monitoring system, in line with global hygiene indicators.

### Evidence-based recommendations

Most recommendations are not supported by evidence (Figure 3). Of the 923 extracted recommendations, only 7% (n= 68) are supported by at least one of the four types of evidence identified for the review. In addition, only 2% (n= 18) are supported by peer-reviewed literature, which are mainly for alternative hand hygiene materials, such as ash, sand, and soil, or alternative water sources for handwashing, such as cooking water, laundry water, bathwater, and seawater. Nonetheless, the cited peer-reviewed literature are individual studies, and not systematic reviews.

**Figure 3.**
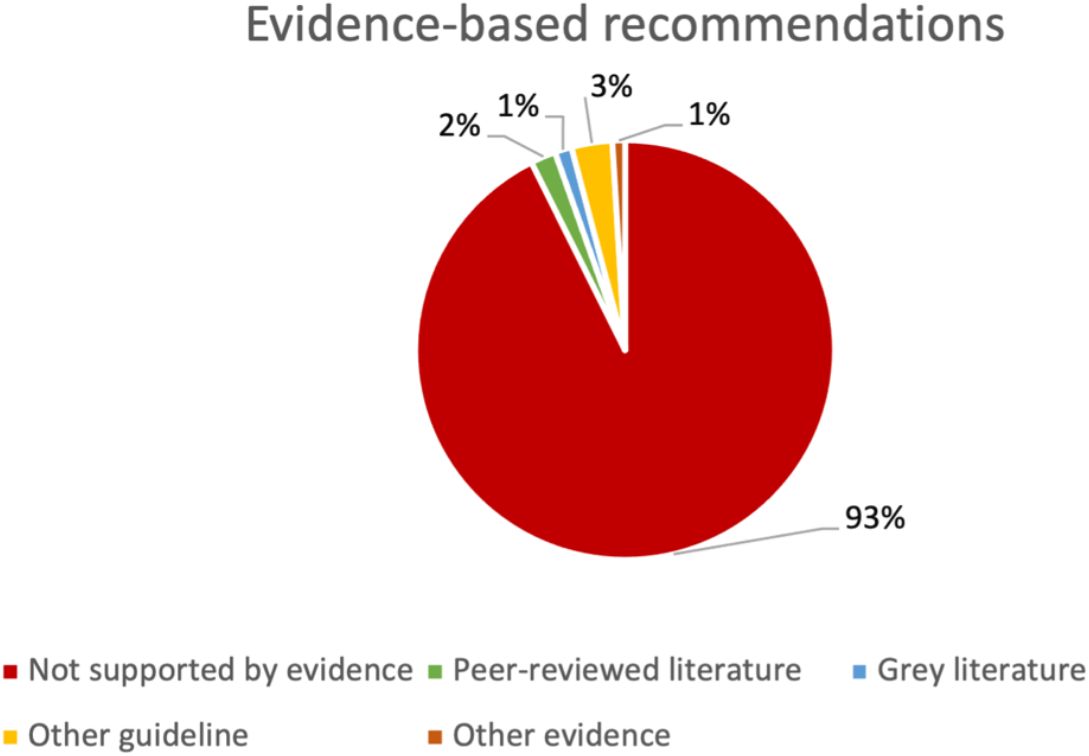
Percentage of recommendations not supported by evidence and by type of evidence.

## DISCUSSION

We identified 51 guidelines published between 1999 and 2021 by various international agencies covering a range of community settings. Most guidelines target the public and institutional setting, while surprisingly none exclusively target the domestic setting. Overall, community settings are not clearly defined among the guidelines, which presents an opportunity for future normative guidelines to establish a clear and common definition, especially as it relates to hand hygiene. Overall, no guidelines comprehensively address hand hygiene across domestic, public, and institutional settings, and very few recommendations are evidence-based, highlighting a gap in global normative guidance on hand hygiene in community settings.

### Effective hand hygiene

There is agreement among guidelines on what constitutes effective hand hygiene, however, inconsistencies and gaps remain as to when and how to practice hand hygiene. Current recommendations for handwashing with soap reflect the literature that suggests that handwashing with soap is an effective means for preventing the transmission of a range of diseases.^3–9^ Regarding when to practice hand hygiene, future guidelines could focus key moments on those most likely to interrupt the transmission of infectious diseases in domestic, public, and institutional settings. These might include after using the toilet to reduce the transmission of diarrhoea-causing pathogens,^9^ and after touching high-contact surfaces or after coughing or sneezing to reduce the risk of respiratory infections.^3^ Lastly, gaps in recommendations on handwashing and hand rubbing technique present an opportunity for future guidelines to recommend the optimal technique for reducing bacterial load on hands.

### Minimum requirements

Consistent recommendations for the use of soap and water and ABHR suggest the widespread acceptability of these materials for effective hand hygiene across community settings. Current recommendations for the use of plain soap are consistent with findings from a systematic review that suggest that plain soap is more effective than antibacterial soap at removing or inactivating pathogens on hands in community settings.^38^ Recommendations for running water are equally relevant as unreliable water supply negatively affects households’ ability to perform hand hygiene.^39^ Gaps in recommendations on minimum quantities of soap and water required for effective hand hygiene are important for future guidelines to address. The recommended use of ABHR with at least 60% alcohol is in line with several studies which have found that ABHRs with an alcohol concentration between 60 and 80% are more effective at killing germs than those with a lower alcohol percentage, particularly in clinical settings.^40,41^ Although ABHR can still inactivate many types of microbes when used correctly, evidence suggests that soap and running water are more effective at removing certain types of pathogens that may be present on hands.^42^ The WHO Guidelines on Hand Hygiene in Health Care, for example, recommend ABHR as the preferred method for routine hand hygiene in healthcare settings when hands are not visibly soiled, as it enables more frequent hand hygiene.^28^ Similarly, in community settings, ABHR may be suitable in contexts where frequent hand hygiene is necessary, such as transport hubs and entrances or exits to public spaces and buildings.^43^ However, in certain domestic, public, and institutional settings, such as the household or schools, where hands may become more soiled, ABHR may be less likely to effectively inactivate microbes.^40,41^ As per current international guidelines, handwashing with soap may therefore be prioritised in these settings, with ABHR as a suitable complement or alternative where frequent hand hygiene is required. Finally, because the transmission of germs is more likely to occur to and from wet hands, hand drying is an essential component of effective hand hygiene, especially for handwashing.^44^ Current recommendations for hand drying are consistent with those in the WHO Guidelines on Hand Hygiene in Health Care, which recommend that hands should ideally be dried with individual paper towels, otherwise with air dryers.

^28^ While there is mixed evidence for the most effective hand drying method,^45^ the WHO recommendations are based on findings that suggest that paper towels may effectively prevent recontamination of hands, while also lowering the risk of spreading pathogens through the air compared to electric air dryers.^44^

Discordant recommendations for minimum requirements suggest the need to leverage further research to determine the optimal water quality for handwashing and effectiveness of alternative materials for hand hygiene where soap and ABHR are not widely available. Limited evidence suggests that the use of non-potable water with low-to-moderate levels of *E. coli* contamination may still be effective for handwashing,^46^ which may be promising for areas where it is difficult to regularly treat water or where there is intermittent water supply that is prone to contamination. Similarly, two studies found that drinkable water may not be needed for handwashing with soap.^47,48^ Nevertheless, the WHO Healthcare Guidelines recommend washing hands with clean, running water whenever possible.^28^ While there are discordant recommendations for the use of ash, there is uncertain evidence whether this stops or reduces the spread of pathogens compared to hand cleansing with soap, mud, soil, or no hand cleansing.^49^ Future guidelines may consider the efficacy of hand hygiene products along with their availability and acceptability in domestic, public, and institutional settings to make relevant recommendations, particularly in water-scarce regions or settings where there is limited access to soap or ABHR. For example, soapy water may be a promising low-cost and effective alternative to bar soap in settings where bar or liquid soap is unaffordable.^50^ One interim guideline on hand hygiene practices in low-resource settings recommends the use of friction-generating materials where clean, running water, soap, or ABHR are not available.^51^

Current inconsistent recommendations for hand hygiene facilities and their placement may limit the practice of effective hand hygiene. Sustaining hand hygiene behaviour change requires consistent access to functional hand hygiene stations at key locations^52,53^ and diverse infrastructure is recommended with varying costs. Nonetheless, the appropriateness of these recommendations is likely to depend on the local availability and affordability of materials. One guideline, for example, provides technical recommendations for permanent and semi-permanent handwashing facilities in public places and buildings, focusing on the sustainability and equitable access of these facilities.^20^ Similarly, future guidelines may prioritise the accessibility, affordability, and sustainability of materials and infrastructure for hand hygiene in domestic, public, and institutional settings to address current inconsistent and discordant recommendations. The accessibility and sustainability of hand hygiene stations are particularly important to ensure that they are inclusive and kept functional and well-stocked beyond their installation.^20^

### Behaviour change

The gaps in recommendations related to behaviour change suggest the need for guidance based on established behavioural theory and existing evidence. There are some recommendations for behaviour change, though without clear steps on how to develop, implement, and sustain hand hygiene behaviour change interventions. Future guidelines may benefit from leveraging well-established behavioural frameworks and theories^54–56^ to make recommendations as to how to develop effective, locally-appropriate strategies beyond information-focused communication. In addition, while most behaviour change theories and frameworks recommended amongst the guidelines note the importance of robust formative research, very few guidelines recommend undertaking formative research. Yet, formative research plays a key role in adapting hand hygiene behaviour change programmes to high-risk populations and target settings.^57^

### Government measures

The significant gaps in recommendations on government measures underscore the current lack of normative standards to guide national governments on the planning, delivery, financing, and monitoring of effective hand hygiene. Indicators also suggest inadequate planning and insufficient funding for hand hygiene among national governments globally.^19^ Future guidelines may therefore consider prioritising government measures to support countries in responding to and preventing public health crises, such as the COVID-19 pandemic. Future guidelines may also focus recommendations on hand hygiene monitoring and reporting to improve comparison of hand hygiene indicators within and between countries.

### Strengths and limitations

This review has three main limitations. First, as a scoping review we did not systematically assess the quality of the included guidelines although we did, for example, consider aspects such as the extent to which recommendations were based on evidence. In addition, included guidelines covered WASH and public health topics beyond hand hygiene, so it was therefore not necessarily relevant to assess the whole guidelines for quality, but rather focus on the robustness of the specific recommendations for hand hygiene. Second, the search was limited to guidelines published in English or French and therefore may have excluded relevant guidelines published in other languages. Third, while recommendations were summarised across domestic, public, and institutional settings, there were often too few recommendations for each setting to assess consistency and concordance. Still, with 51 guidelines providing over 900 recommendations for hand hygiene in community settings, findings from this review highlight significant gaps and inconsistencies that future guidelines may seek to prioritise.

## CONCLUSION

This review identified 51 current international guidelines for hand hygiene in community settings providing 923 recommendations. Nonetheless, there are several important areas of discordance and significant gaps in the recommendations among these guidelines. Furthermore, very few recommendations are supported by any qualifying evidence. The COVID-19 pandemic led to numerous national, regional, and international efforts to improve effective hand hygiene in domestic, public, and institutional settings, such as households, public spaces, workplaces, and schools, but the lack of clear, evidence-based recommendations may limit progress in this important area of public health.

## Data Availability

All data produced in the present work are contained in the manuscript

## Contributors

CM, LB, BC, CC, KC, JC, RD, REN, BG, JEM, and OC informed the study protocol. CM carried out the database and grey literature search with input from OC and JEM. JEM, OC, BG, and CM carried out expert consultations. CM and LB screened the retrieved articles for inclusion and extracted the data with input from OC. CM led on data analysis and CM, LB, OC, and JEM led on the presentation of results with inputs from co-authors. CM, LB, JEM, and OC led on writing the manuscript with input from all co-authors. OC, JEM, and BG provided overall supervision, leadership, and advice. The first author (CM) accepts responsibility for the work.

## Patient and public involvement

Patients and/or the public were not involved in the design, or conduct, or reporting, or dissemination plans of this research.

## Patient consent for publication

Not applicable.

## Ethics approval

This research did not require institutional review board approval as the data were publicly available and collected from existing online databases and search engines. This research did not involve any human subjects.

## Data availability statement

All data relevant to the study are included in the article or uploaded as supplementary information.

