## Supplementary material for "Improving hand hygiene in community settings: a scoping review of current international guidelines"

**SUPPLEMENTAL MATERIALS**

**Table A1**. The completed Preferred Reporting Items for Systematic reviews and Meta-Analyses extension for Scoping Reviews (PRISMA-ScR) checklist.[25]

| SECTION | ITEM | PRISMA-ScR CHECKLIST ITEM | REPORTED ON PAGE # |
|---|---|---|---|
| **TITLE** | | | |
| Title | 1 | Identify the report as a scoping review. | 2 |
| **ABSTRACT** | | | |
| Structured summary | 2 | Provide a structured summary that includes (as applicable): background, objectives, eligibility criteria, sources of evidence, charting methods, results, and conclusions that relate to the review questions and objectives. | 3 |
| **INTRODUCTION** | | | |
| Rationale | 3 | Describe the rationale for the review in the context of what is already known. Explain why the review questions/objectives lend themselves to a scoping review approach. | 5 |
| Objectives | 4 | Provide an explicit statement of the questions and objectives being addressed with reference to their key elements (e.g., population or participants, concepts, and context) or other relevant key elements used to conceptualize the review questions and/or objectives. | 5 |
| **METHODS** | | | |
| Protocol and registration | 5 | Indicate whether a review protocol exists; state if and where it can be accessed (e.g., a Web address); and if available, provide registration information, including the registration number. | 5 |
| Eligibility criteria | 6 | Specify characteristics of the sources of evidence used as eligibility criteria (e.g., years considered, language, and publication status), and provide a rationale. | 6 |
| Information sources | 7 | Describe all information sources in the search (e.g., databases with dates of coverage and contact with authors to identify additional sources), as well as the date the most recent search was executed. | 6 |
| Search | 8 | Present the full electronic search strategy for at least 1 database, including any limits used, such that it could be repeated. | Supplemental material Table A5 |
| Selection of sources of evidence | 9 | State the process for selecting sources of evidence (i.e., screening and eligibility) included in the scoping review. | 6 |
| Data charting process | 10 | Describe the methods of charting data from the included sources of evidence (e.g., calibrated forms or forms that have been tested by the team before their use, and whether data charting was done independently or in duplicate) and any processes for obtaining and confirming data from investigators. | 6 |
| Data items | 11 | List and define all variables for which data were sought and any assumptions and simplifications made. | 6 |
| Critical appraisal of individual sources of evidence | 12 | If done, provide a rationale for conducting a critical appraisal of included sources of evidence; describe the methods used and how this information was used in any data synthesis (if appropriate). | 6 – 7 |

| SECTION | ITEM | PRISMA-ScR CHECKLIST ITEM | REPORTED ON PAGE # |
|---|---|---|---|
| Synthesis of results | 13 | Describe the methods of handling and summarizing the data that were charted. | 6 – 7 |
| **RESULTS** | | | |
| Selection of sources of evidence | 14 | Give numbers of sources of evidence screened, assessed for eligibility, and included in the review, with reasons for exclusions at each stage, ideally using a flow diagram. | 7 |
| Characteristics of sources of evidence | 15 | For each source of evidence, present characteristics for which data were charted and provide the citations. | Supplemental material Table A6 |
| Critical appraisal within sources of evidence | 16 | If done, present data on critical appraisal of included sources of evidence (see item 12). | 14 |
| Results of individual sources of evidence | 17 | For each included source of evidence, present the relevant data that were charted that relate to the review questions and objectives. | 7 – 14 |
| Synthesis of results | 18 | Summarize and/or present the charting results as they relate to the review questions and objectives. | 7 – 14 |
| **DISCUSSION** | | | |
| Summary of evidence | 19 | Summarize the main results (including an overview of concepts, themes, and types of evidence available), link to the review questions and objectives, and consider the relevance to key groups. | 14 – 16 |
| Limitations | 20 | Discuss the limitations of the scoping review process. | 16 |
| Conclusions | 21 | Provide a general interpretation of the results with respect to the review questions and objectives, as well as potential implications and/or next steps. | 6 |
| **FUNDING** | | | |
| Funding | 22 | Describe sources of funding for the included sources of evidence, as well as sources of funding for the scoping review. Describe the role of the funders of the scoping review. | 17 |

**Table A2**. List of websites searched.

| Agency or resource hub | Website |
|---|---|
| Africa Centers for Disease Control and Prevention (CDC) | https://africacdc.org/ |
| Global Handwashing Partnership | https://globalhandwashing.org/ |
| Global WASH Cluster | www.washcluster.org |
| International Red Cross and Red Crescent (ICRC) | www.icrc.org |
| International Federation of the Red Cross (IFRC) | www.ifrc.org |
| International Labour Organization (ILO) | https://www.ilo.org |
| International Centre for Diarrhoeal Disease Research Bangladesh (ICDDR'B) | www.icddrb.org |
| Oxfam | www.oxfam.org.uk |

| | |
|---|---|
| Sanitation Learning Hub | https://sanitationlearninghub.org/ |
| Save the Children | www.nrc.no |
| SNV Netherlands Development Organisation | https://snv.org/ |
| Sustainable Sanitation Alliance | https://www.susana.org/en/ |
| United Nations Human Settlement Programme (UN Habitat) | https://unhabitat.org/ |
| United Nations High Commissioner for Refugees (UNHCR) | www.unhcr.org |
| United Nations Children's Fund (UNICEF) | www.unicef.org |
| US Centers for Disease Control and Prevention (CDC) | www.cdc.gov |
| WASH'Em | https://www.washem.info/ |
| WaterAid | https://www.wateraid.org/us/ |
| World Bank | https://www.worldbank.org/en/home |
| Water, Education and Development Centre (WEDC) | www.wedc.lboro.ac.uk |
| Water and Sanitation for the Urban Poor (WSUP) | https://www.wsup.com/ |
| Deutsche Gesellschaft für Internationale Zusammenarbeit (GIZ) | https://www.giz.de |
| United States Agency for International Development (USAID) | https://www.usaid.gov/ |

**Table A3**. Search terms.

| Concept | Search terms |
|---|---|
| Hand hygiene | Handwash* OR hygiene OR hand wash* OR with soap OR hand hygiene* OR clean hands OR WASH |
| Community settings | community OR household OR domestic OR public space OR institution* OR work* OR occupation* OR school OR prison OR market* OR relig* OR education* OR detention* |
| Guideline | guideline OR guide OR handbook OR manual OR toolkit |

**Table A4**. Hand Hygiene for All Initiative core partners**.**

| |
|---|
| United Nations Children's Fund |
| International Labour Organization |
| World Bank |
| United Nations High Commission for Refugees |
| COVID-19 Hygiene Hub |
| Sanitation and Water for All |
| Global Handwashing Partnership |
| International Federation of Red Cross |
| WaterAid |

**Table A5**. Excluded documents with reasons.

| Author | Guideline title | Year | Reason for exclusion |
| --- | --- | --- | --- |
| ICRC | Water, sanitation, and hygiene, and habitat in prisons | 2005 | Historical version of guideline |
| World Bank | The Handwashing Handbook: a guide for developing a hygiene promotion program to increase handwashing with soap | 2005 | Historical version of guideline |
| IRC | Strengthening Water, Sanitation and Hygiene in schools: a WASH guidance manual with a focus on South Asia | 2010 | Not a guideline |
| ICRC | Water, sanitation, and hygiene, and habitat in prisons: supplementary guidance | 2012 | No recommendation on hand hygiene |
| GIZ | Field guide: hardware for handwashing in schools | 2013 | Not a guideline |
| WEDC | Guidelines for handwashing with soap | 2013 | Not issued by an international agency |
| NRC | Water, sanitation and hygiene manual: WASH training for hygiene promotion staff | 2015 | Not a guideline |
| ILO | WASH@Work: self-training handbook | 2016 | Historical version of guideline |
| WSUP | School hygiene manual | 2016 | Not for international audience |
| Rotary | A guide to WASH in schools | 2017 | Not a guideline |
| Global Handwashing Partnership | Clean hands for all: a toolkit for hygiene advocacy | 2018 | Not a guideline |
| COOPI | WASH handbook for teachers and facilitators | 2019 | Not for international audience |
| WASH'Em | How to design handwashing facilities that change behaviour | 2019 | Does not target a community setting |
| Africa CDC | Hand washing facility options for resource limited settings | 2020 | Not a guideline |
| British Psychological Society | Behavioural science and disease prevention psychological guidance: encouraging hand hygiene in the community | 2020 | Not a guideline |
| Coultas et al. | Handwashing compendium for low-resource settings: a living document, 3rd edition | 2020 | Not a guideline |
| Global Handwashing Partnership | The Handwashing Handbook | 2020 | Not a guideline |
| Smart Centre Group | Smart hygiene solutions: affordable options for household level | 2020 | Not a guideline |

| | | | |
|---|---|---|---|
| UN Habitat | UN-Habitat guidance on COVID-19 and public spaces | 2020 | No recommendation on hand hygiene |
| UNHCR | COVID-19 refugees return to schooling guidelines | 2020 | No recommendation on hand hygiene |
| UNICEF et al. | Panduan Cuci Tangan Pakai Sabun | 2020 | Not in English or French |
| WaterAid | COVID-19 guidance: prioritising hygiene for workforce health and business resilience | 2020 | No recommendation on hand hygiene |
| WaterAid Nepal | Technical brief on contactless handwashing stations | 2020 | Not a guideline |
| Bolton, L. | WASH in schools for student return during the COVID-19 pandemic | 2021 | Not issued by an international agency |
| ILO | WASH@Work: self-training handbook (revised) | 2021 | Not a guideline |
| Government of Vietnam | A guide to integrated hand washing with soap communication | n/d | Not for international audience |
| WaterAid | Technical manual on community water supply, hygiene and sanitation facilities | n/d | Not a guideline |
| WaterAid Bangladesh | Handwashing stations: an easy-to-use technological and context-based handwashing stations manual | n/d | Not a guideline |

**Table A6**. List of included guidelines.

| Author | Guideline | Year | Community setting | COVID-19 response |
|---|---|---|---|---|
| UNICEF | A manual on hygiene promotion | 1999 | Public | No |
| WHO | WHO recommended strategies for the prevention and control of communicable diseases | 2001 | Domestic & public | No |
| WHO | Health villages: a guide for communities and community health workers | 2002 | Public | No |
| WHO | Water, sanitation, and hygiene standards for schools in low-cost settings | 2009 | Institutional | No |
| ICRC | Water, sanitation, hygiene, and habitat in prisons | 2013 | Institutional | No |
| ICRC | Health care in detention | 2015 | Institutional | No |
| WHO | Public health for mass gatherings: key considerations | 2015 | Public | No |
| SNV | Behaviour change communication guidelines | 2016 | Public | No |
| UNICEF, GIZ | Scaling up group handwashing in schools: compendium of group washing facilities across the globe | 2016 | Schools | No |
| Malteser | WASH guidelines for field practitioners | 2017 | Multiple settings | No |
| Oxfam | Handwashing technical briefing note | 2017 | Public | No |

| | | | | |
|---|---|---|---|---|
| UNHCR | UNHCR hygiene promotion guidelines | 2017 | Public | No |
| WHO | Guidelines on sanitation and health | 2018 | Domestic & public | No |
| WHO | Recommendations to Member States to improve hand hygiene practices to help prevent COVID-19 | 2020 | Public | Yes |
| UNICEF | Handwashing stations and supplies for the COVID-19 response | 2020 | Domestic & public | Yes |
| ILO | Hand hygiene at the workplace: an essential occupational safety and health prevention and control measure against COVID-19 | 2020 | Workplace | Yes |
| GOAL | WASH and IPC measures in households and public spaces | 2020 | Domestic & public | No |
| WHO | Considerations for community hand hygiene practices in low-resource situations | 2020 | Public | No |
| WHO | Overview of public health and social measures in the context of COVID-19 | 2020 | Public | Yes |
| GWC | COVID-19 response guidance note - 15 April 2020 | 2020 | Domestic & public | Yes |
| GWC | Health and hygiene promotion guidance document for COVID-19 | 2020 | Public | Yes |
| ILO | Safe return to work: guide for employers on COVID-19 prevention | 2020 | Institutional | Yes |
| SNV | Practical options for hand-washing stations: a guide for promoters and producers | 2020 | Multiple settings | No |
| UNHCR | Technical WASH guidance for COVID-19 preparedness and response | 2020 | Public | Yes |
| UNICEF | COVID-19 emergency preparedness and response: WASH and infection prevention and control measures in schools | 2020 | Schools | Yes |
| UNICEF | COVID-10 handwashing with soap (HWWS) facilities: compendium of indicative layouts, designs, and cost estimates | 2020 | Public | Yes |
| UNICEF et al. | Handwashing with soap in schools: a technical options manual | 2020 | Schools | No |
| UNICEF, WHO, IFRC | Key messages and actions for COVID-19 prevention and control in schools | 2020 | Schools | Yes |
| UNICEF | Understanding Hygiene promotion in the context of Risk Communication & Community Engagement (RCCE) and Infection Control and Prevention (IPC) for the COVID-19 outbreak | 2020 | Domestic & public | Yes |
| UNICEF | WASH and Infection Prevention and Control (IPC) measures in households and public spaces | 2020 | Domestic & public | Yes |
| UNICEF, WHO, IFRC | Interim guidance for COVID-19 prevention and control in schools | 2020 | Institutional | Yes |
| WASH4WORK | Hand hygiene protocol for the workplace | 2020 | Institutional | Yes |
| Water Mission | Handwashing station program: requirements and recommendations | 2020 | Public | Yes |
| WaterAid | Technical guide for handwashing facilities in public spaces and buildings | 2020 | Public | Yes |
| WHO | Actions for consideration in the case and protection of vulnerable populations from COVID-19 | 2020 | Vulnerable populations | Yes |

| | | | | |
|---|---|---|---|---|
| WHO | Considerations for public health and social measures in the workplace in the context of COVID-19: Annex to Considerations in adjusting public health and social measures in the context of COVID-19 | 2020 | Workplace | Yes |
| WHO | Considerations for school-related public health measures in the context of COVID-19: Annex to Considerations in adjusting public health and social measures in the context of COVID-19 | 2020 | Institutional | Yes |
| WHO | Guidance on COVID-19 for the care of older people and people living in long-term care facilities, other non- acute care facilities and home care | 2020 | Institutional | Yes |
| WHO | Interim recommendations on obligatory hand hygiene against transmission of COVID-19 | 2020 | Public spaces | Yes |
| WHO | Practical considerations and recommendations for religious leaders and faith-based communities in the context of COVID-19 | 2020 | Public spaces | Yes |
| WHO | Water, sanitation, hygiene, and waste management for SARS-CoV-2, the virus that causes COVID-19: interim guidance | 2020 | Domestic & public | Yes |
| CDC | When and how to wash your hands | 2021 | Public | No |
| CDC | Hand hygiene at school | 2021 | Institutional | No |
| CDC | Hand hygiene at work | 2021 | Institutional | No |
| CDC | Interim guidance on management of coronavirus disease 2019 (COVID-19) in correctional and detention facilities | 2021 | Institutional | Yes |
| CDC | Interim guidance on people experiencing unsheltered homelessness | 2021 | Vulnerable populations | Yes |
| WHO | Key planning recommendations for mass gatherings in the context of COVID-19: interim guidance | 2021 | Public | Yes |
| WHO | Preparedness, prevention, and control of COVID-19 in prisons and other places of detention: interim guidance | 2021 | Institutional | Yes |
| WHO, ILO | Preventing and mitigating COVID-19 at work | 2021 | Institutional | Yes |
| WHO | Safe Eid al Adha practices in the context of COVID-19: interim guidance | 2021 | Public | Yes |
| WHO | Safe Ramadan practices in the context of COVID-19: interim guidance | 2021 | Public | Yes |

**Table A7**. Number of recommendations per parameter across all community settings.

| Parameters | Number of recommendations | Number of guidelines with recommendations |
|---|---|---|
| **Safe hand hygiene** | | |
| Guidelines that define hand hygiene | 3 | 3 |
| Guidelines that define safe hand hygiene | 2 | 2 |
| Critical control points language | 38 | 38 |
| Critical control points | 145 | 38 |
| Basic handwashing knowledge | 8 | 8 |
| Basic hand hygiene knowledge | 0 | 0 |
| **Handwashing with soap & hand drying** | | |
| Handwashing infrastructure | 33 | 24 |
| Handwashing materials | 68 | 46 |
| Amount of soap | 7 | 6 |
| Handwashing services | 37 | 30 |
| Handwashing with soap duration | 17 | 17 |
| Water quality | 19 | 16 |
| Water quantity | 9 | 10 |
| Alternative water sources for handwashing | 15 | 16 |
| Greywater management | 23 | 13 |
| Location of handwashing stations | 59 | 30 |
| Spacing and number of users per handwashing station | 9 | 8 |
| Hand drying materials | 36 | 19 |
| Disposal of hand drying materials | 9 | 9 |
| Conditional recommendations | 7 | 7 |
| **Handwashing with soap alternatives** | | |
| Alcohol-based hand rub | 32 | 32 |
| Hand rubbing duration | 3 | 3 |
| Other materials for hand hygiene | 39 | 17 |
| Conditional recommendations | 29 | 22 |
| Alternatives not recommended | 11 | 7 |
| Location of hand hygiene stations | 21 | 12 |
| **Hand hygiene station design considerations** | | |
| Design principles | 10 | 6 |
| Technical considerations | 5 | 4 |
| Accessibility | 19 | 14 |
| Local procurement and production | 11 | 9 |
| Supply chain | 3 | 3 |
| Cost and affordability | 7 | 7 |
| Durability | 8 | 6 |
| COVID-19 adaptations | 26 | 16 |
| Other adaptations | 17 | 10 |
| Operation and maintenance – responsibility | 8 | 6 |
| Operation and maintenance – actions | 9 | 8 |
| **Changing behaviour** | | |
| Behaviour change techniques | 29 | 14 |
| Content of behaviour change messaging | 27 | 25 |
| Behaviour change delivery channels | 42 | 25 |
| Person or group tasked with delivery behaviour change intervention | 13 | 10 |
| Frequency of behaviour change interventions | 2 | 2 |
| Formative research | 5 | 4 |

| | | |
|---|---|---|
| Behaviour change models or frameworks | 15 | 3 |
| Behaviour change approaches | 9 | 2 |
| Hand hygiene determinants | 11 | 3 |
| **Government interventions** | | |
| Strengthening markets for hand hygiene products and services | 2 | 2 |
| Hygiene promotion practices | 0 | 0 |
| At-scale behaviour change interventions | 0 | 0 |
| Sector policy and strategy | 0 | 0 |
| Institutional arrangements | 5 | 5 |
| Sector financing | 0 | 0 |
| Planning, monitoring, and review | 2 | 2 |
| Capacity development | 1 | 1 |
| Equity | 1 | 1 |
